# How Policies on Restaurants, Bars, Nightclubs, Masks, Schools, and Travel Influenced Swiss COVID-19 Reproduction Ratios

**DOI:** 10.1101/2020.10.11.20210641

**Authors:** C. K. Sruthi, Malay Ranjan Biswal, Brijesh Saraswat, Himanshu Joshi, Meher K. Prakash

## Abstract

The role of complete lockdowns in reducing the reproduction ratios (R_t_) of COVID-19 is now established. However, the persisting reality in many countries is no longer a complete lockdown, but restrictions of varying degrees using different choices of Non-pharmaceutical interaction (NPI) policies. A scientific basis for understanding the effectiveness of these graded NPI policies in reducing the R_t_ is urgently needed to address the concerns on personal liberties and economic activities. In this work, we develop a systematic relation between the degrees of NPIs implemented by the 26 cantons in Switzerland during March 9 – September 13 and their respective contributions to the R_t_. Using a machine learning framework, we find that R_t_ which should ideally be lower than 1.0, has significant contributions in the post-lockdown scenario from the different activities - restaurants (0.0523 (CI. 0.0517-0.0528)), bars (0.030 (CI. 0.029-0.030)), and nightclubs (0.154 (CI. 0.154-0.156)). Activities which keep the land-borders open (0.177 (CI. 0.175-0.178)), and tourism related activities contributed comparably 0.177 (CI. 0.175-0.178). However, international flights with a quarantine did not add further to the R_t_ of the cantons. The requirement of masks in public transport and secondary schools contributed to an overall 0.025 (CI. 0.018-0.030) reduction in R_t_, compared to the baseline usage even when there are no mandates. Although causal relations are not guaranteed by the model framework, it nevertheless provides a fine-grained justification for the relative merits of choice and the degree of the NPIs and a data-driven strategy for mitigating R_t_.

## 1. Introduction

COVID-19 has caused over a million deaths, and many millions of infections globally. Despite the intense global efforts, there is no scalable treatment or vaccine yet. This problem is compounded by the fact that the epidemiology of COVID-19 is very unique [2] and that it spreads efficiently, with an estimated 44% [3] to 69% [4] of the transmissions before the onset of symptoms. Non-pharmaceutical interventions (NPI) have been the only successful method of mitigating the spread of infections [5-7]. In addition to the initial recommendations on social-distancing and sanitizing, complete lockdowns and restricted opening of businesses had been implemented in various degrees. There have been several demonstrations of the effectiveness of the lockdowns in successfully reducing the reproduction ratio (R_t_) and mitigating the spread of COVID-19 [8-11].

NPIs performing a balancing act. The goal of NPIs is to flatten the infection curve [12] until there is a change in the infection landscape due to a scalable vaccine, therapeutic intervention, or a mutation in the virus which reduces the case fatality rate. However, since NPIs affect both personal liberties and economic welfare, it is important to know the effectiveness of the different NPIs for evidence-based public health planning. The impact of the two extremes of the NPI spectrum [8-11], no restrictive measures and a complete lockdown are relatively easy to understand. However, many policies changes, such as the compulsory use of masks, different degrees of opening of the restaurants or bars have been applied simultaneously, and it has not been easy to quantify the effectiveness of the contributions from these different factors. To the best of our knowledge, there have not been any examples of deciphering the contributions of the policies to the R_t_.

In this work, we apply a recently developed formalism [13] to decipher the role of the different NPI policies adopted by Switzerland. Until April, Switzerland had the highest per-capita COVID-19 infections globally, however Switzerland could open up most of its activities staring May. During May-July period, the number daily new infections was less than 100, and it was followed by increasing infections as well as tightening of regulations. The few cases together with an efficient infrastructure for contact tracing [14] helped isolate all potentially infected persons, and helped in mitigating the spread of infections. However, as the daily new cases crossed 1,000 the fears that the efficiency of the contact tracing are reaching the limits begins to resound. Further, another complicated aspect of the epidemiology of COVID-19 has been that the infections need not always be attributable to a symptomatic person. The ability to understand the overall impact of an NPI policy on the overall infections, which implicitly includes the secondary infections, asymptomatic transmissions is thus needed and is the focus of the present article.

## 2. Results

### 2.1 Relating the policies to reproduction ratios

#### 2.1.1 Determinants - NPI Policy data

To perform a quantitative analysis, the different degrees of non-pharmaceutical intervention policies were curated and codified. Media reports published in english language were used for this curation and the policies were cross-checked with the official press releases of the Federal and Cantonal authorities. The following codes were employed:

- *BarbersSalonSpa*: Everything closed (0), Beauty salons, Barbers shops etc allowed to operate with the stipulated norms (1), Pre-lockdown level (2)
- *Bars*: Closed (0), Opened with limited capacity and restrictions on standing customers (1), Opened with full capacity while respecting social distancing (2), Pre-lockdown level (3)
- *Mask*: No mask requirement (0), Mask usage is mandatory in public transport (1), Mask usage is mandatory in shops (2)
- *Nightclubs and Discos*: Closed (0), Opened with contact tracing and checking the ID or with a limited audience of 50 to 100 (1), Opened with an audience of over 100 up to 300 (2), Pre-lockdown level (3)
- *Restaurants*: Closed (0), Takeaway only (1), restaurant occupancy limited to 50 (2), restaurant occupancy of 100 (3), Unrestricted but respecting social distancing (4), Pre-lockdown level (5)
- *Retail*: All non-essentials closed (0), Opening do-it stores (1), Opening all retail stores (2), Pre-lockdown level (3)
- *Schools*. Closed (0), Open with mask-requirements on pupils in all secondary-schools or when the distance between pupils is less than 2.25 meters (1), Open with no mask-requirements
- *Tourism potential*. The number of hotel rooms available in the cantons, invariable with time. This can be a surrogate for the general mobility and associated economic activity in the regions
- *Travel restrictions*: All travel from outside Switzerland is restricted (0), Travel by road to the cantons having a road-entry to Schengen countries is opened (1), In addition to the road travel from Schengen countries, International travel is permitted with a quarantine requirement upon arrival (2)

In this work we use the NPIs and R_t_’s with the resolution of a week [15]. Each NPI policy code was assumed to be the same for an entire week starting from Monday to Sunday. The data was applied over 26 weeks between March 9 to September 13. Several exceptional cases have not been included in the choice of the policy variables. Manufacturing was shutdown specifically in the early lockdown phase in Ticino (TI) canton. The states in USA had two major differences – “stay at home” versus “safe at home”. The Swiss policies reflected more the latter, and unlike some states of USA or Italy had not restricted outdoor activities. Hence the permission for outdoor activities was not included in the analysis.

#### 2.1.2 Consequences - Reproduction rates (R_t_)

Time varying reproduction rate, R_t_ (=*α*_t_ /*γ*) was estimated using the rates of transmission (*α*_t_) and recovery from the infection (*γ*). The infected and recovered data was obtained from *https://www.corona-data.ch/*. We estimated the average recovery time in for Switzerland to be around 14 days, and *γ*=1/14. Using the number of active infections (I_active_), and the number of daily new infections on a given day (dI/dt), the weekly averaged transmission rates were calculated as, *α*_t_=<1/I_active_. (dI/dt)>.

#### 2.1.3 Model - Interpretable Artificial Intelligence

Our goal is to develop a model relating the controlling NPIs and the consequent reproduction ratios by a model, and to understand the relative contributions of the NPIs to the R_t_. The NPIs of any week are assumed to affect the reproduction rate in the following week. To not restrict the model to linearity and maintain the generality, we used an artificial intelligence model. The data from 26 weeks and 26 cantons used, with 20% for testing and 80% for training. A 5-fold cross validation was used to further validate the results. The daily rates, their weekly averages *α*_t_ and how they change over the weeks is illustrated in Figure 2 and demonstrates a dynamic scenario.

**Figure 1.**
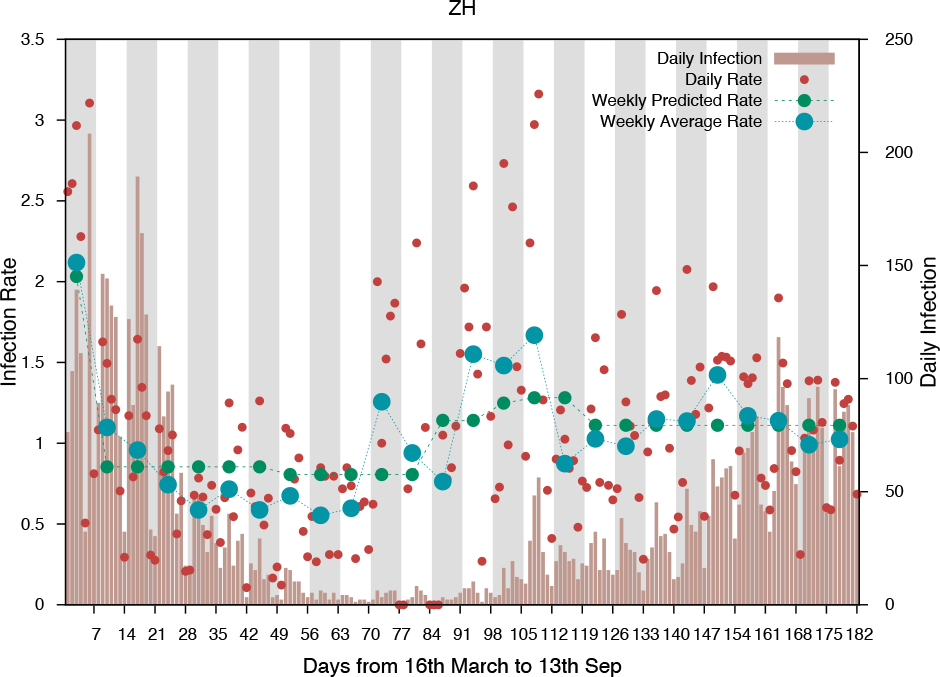
Comparing the observations and estimations from Zurich canton. The number of daily infection cases, the daily instantaneous R_t_, and its weekly average are shown. Also shown is the R_t_ predicted using our AI model. The quality of predictions for the different cantons are shown in Table 1.

**Figure 2.**
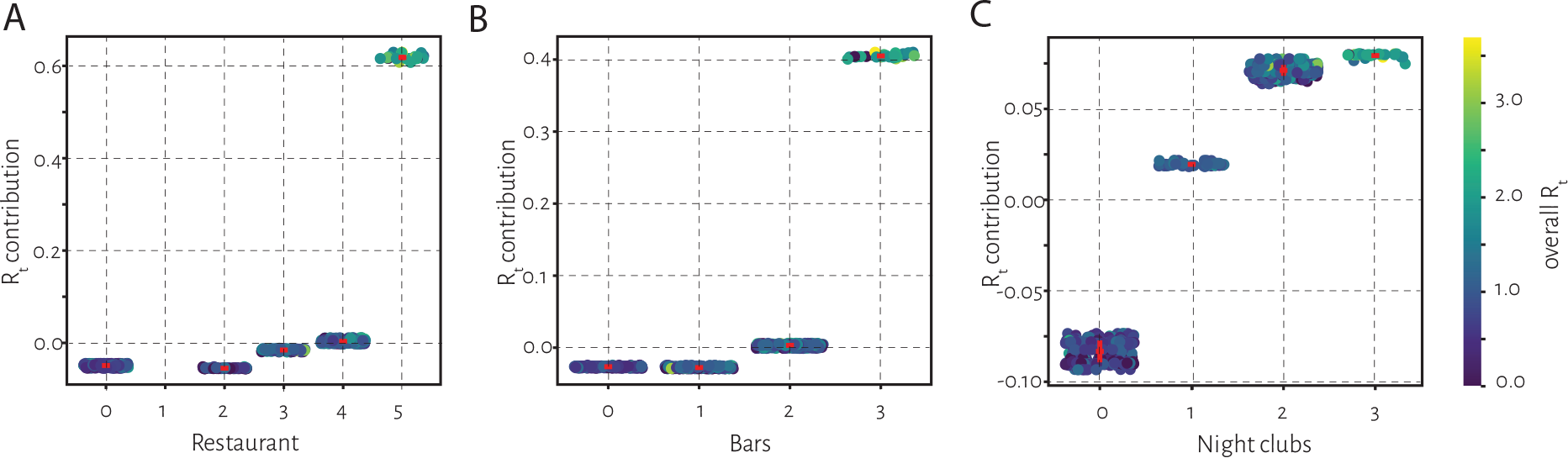
Impact of restrictive measures on dine and drink. The contributions to the R_t_ from the different restrictive policies for operating *A*. Restaurants *B*. Bars and *C*. Nightclubs are shown. Each point in the three graphs represents the contribution to the R_t_ from a specific week in one of the 26 cantons, and the colorbar represents the value of the R_t_. In all three graphs, 0 on the x-axis represents the complete closure, and the highest value the pre-lockdown operational levels. The red lines indicate the confidence interval in each category.

The artificial intelligence (AI) model was developed using the *XGBoost* library in Python. 80% of the complete data set was used as the training set and 20% as the test set. The parameters for the AI model (*learning_rate*=0.08, *n_estimators*=525, *max_depth*=7, *min_child_weight*=5, *gamma=*0, *subsample*=0.85, *colsample_bytree*=0.75, *reg_alpha*=1) were obtained using *RandomizedSearchCV*. With these parameters, the predictions of the reproduction rates for the Cantons were good as shown in Table 1. Each model’s predictive ability is evaluated using a 5-fold cross-validation analysis and the best set of parameters is selected based on the average of mean squared error for the validation sets. The quality of prediction seemed to be robust against changes in the training set as shown by the 5-fold cross-validation analysis.

**Table 1.** Quality of predictions for the cantons. The co-efficient of determination R^2^ is used as a metric to understand if the model results, including training and test, compare well with the R_t_ estimates. The results are good when the number of cases recorded were more than 1000. The uncertainties in both the estimation of R_t_ from the epidemiological data, and the predictions using a model are both expected when the number of infections are fewer.

| Canton | Cases until 13 September | $R^2$ from a single model | $R^2$ from 5-fold Cross Validation | Canton | Cases until 13 September | $R^2$ from a single model | $R^2$ from 5-fold Cross Validation |
| --- | --- | --- | --- | --- | --- | --- | --- |
| VD | 8795 | 0.673117037 | 0.647847665 | NE | 985 | 0.25887226 | 0.21571116 |
| GE | 7616 | 0.627418522 | 0.625777595 | SO | 744 | 0.210001297 | 0.187173494 |
| ZH | 7008 | 0.598277452 | 0.586055896 | TG | 640 | 0.389502291 | 0.410028348 |
| TI | 3577 | 0.29952398 | 0.283008382 | SZ | 527 | -0.07411167 | -0.126575857 |
| BE | 2852 | 0.368545109 | 0.379780423 | ZG | 392 | 0.247699512 | 0.199368169 |
| VS | 2505 | 0.459523001 | 0.449849909 | JU | 289 | 0.137647876 | 0.120101292 |
| AG | 2364 | 0.618817663 | 0.640935414 | SH | 177 | 0.369710129 | 0.369761684 |
| FR | 2106 | 0.431110605 | 0.387268328 | GL | 168 | 0.140084061 | 0.13367399 |
| SG | 1414 | 0.384846143 | 0.374026311 | AR | 139 | -0.01352006 | -0.021623142 |
| BS | 1272 | 0.236721344 | 0.243603321 | NW | 139 | 0.035126862 | 0.068942698 |
| BL | 1137 | 0.341130146 | 0.33396309 | UR | 138 | 0.146536746 | 0.172496426 |
| LU | 1115 | 0.535569715 | 0.534524372 | OW | 108 | -0.21602673 | -0.20605954 |
| GR | 1049 | 0.311370476 | 0.328251839 | AI | 26 | -0.67085668 | -0.49664838 |

### 2.2 Decoupling the contributions from the different NPIs

#### 2.2.1 Interpretable AI for relative contributions

We performed a retrospective analysis to understand the relation between the different non-pharmaceutical interventions policies imposed by each canton in every week between March 9 to September 13, which we codified as discussed above, and the reproduction ratios (R_t_). To decouple the relative contributions from the individual NPIs to each of the weekly predictions, we post-processed the results of our AI model using an interpretable AI framework based on SHapley Adaptive exPlanation (SHAP) [16]. A Python implementation of SHAP (https://github.com/slundberg/shap) was used for our interpretable AI analyses. The SHAP analyses was repeated with a 5-Fold Cross-Validation and the variances were found to be minimal. The role of many NPIs with significant contributions to R_t_ are discussed below.

#### 2.2.2 Dine and drink

The contributions to R_t_ from the different graded openings of the restaurants is shown in Figure 2A. The x-axis represents the different grades of opening of the restaurants: restaurants are closed (0), open with up to 50 clients (2), open with up to 100 clients (3), unrestricted clientele but following social distancing norms (4), pre-lockdown level (5). The restaurant-component to R_t_ from the predictions for the 26 cantons and 26 weeks (y-axis) is organized by the restaurant opening degree and displayed. The color of these points was used to represent the overall R_t_ in that canton in that week. The data shows a clear pattern of increasing contributions to the R_t_ from the increasing degrees of restaurant openings. Leaving the pre-lockdown conditions (code 5), which lacked sanitization, social-distancing measures, the maximum R_t_ one can expect because of opening restaurants at level 4 relative to a complete closure is 0.0523 (CI. 0.0517-0.0528). Similar analysis shows that different graded openings of the bars (Figure 2B) between a full opening of the bars (code 2) a complete closure (code 0) contribute 0.030 (CI. 0.029-0.030) to R_t_. Nightclubs (Figure 2C) with more than 100 clients allowed (code 3) contributes 0.154 (CI. 0.154-0.156) to the R_t_ relative to their complete closure, and is the only case in all of our analysis where the R_t_ contribution is comparable to the contributions from the pre-lockdown period.

#### 2.2.3 Travel and tourism

In March, Switzerland had closed entry access via road to Schengen countries as well as via air travel. Starting from mid-June, the road access to Schengen countries was opened (code 1 for cantons sharing borders with neighboring countries) and air-travel was permitted with a quarantine requirement upon arrival (code 2 for cantons with international airports). The contributions to R_t_ from this ease-of-access descriptor shown in Figure 3A, shows an additional R_t_ of 0.177 (CI. 0.175-0.178) due to opening of the land-borders. The international air-travel with a quarantine requirement did not contribute any further to the R_t_. An additional factor that was included in the analysis was the tourism potential quantified by the number of hotel rooms in the canton, which of course remains constant during the 26 weeks of analysis. The variable contributed 0.179 (CI. 0.172-0.187) to the R_t_. The tourism potential is neither the actual number of travels made in this year, nor an NPI policy, but a factor which remained a constant for each canton. However, given the magnitude of its contribution, it requires an interpretation, possibly of the surrogate it represents. Specifically, the cantons JU (code 102), AR (code 122) which have lower number of hotels are near the borders and have a high cross-border commuter traffic which is probably being captured in the analysis.

**Figure 3.**
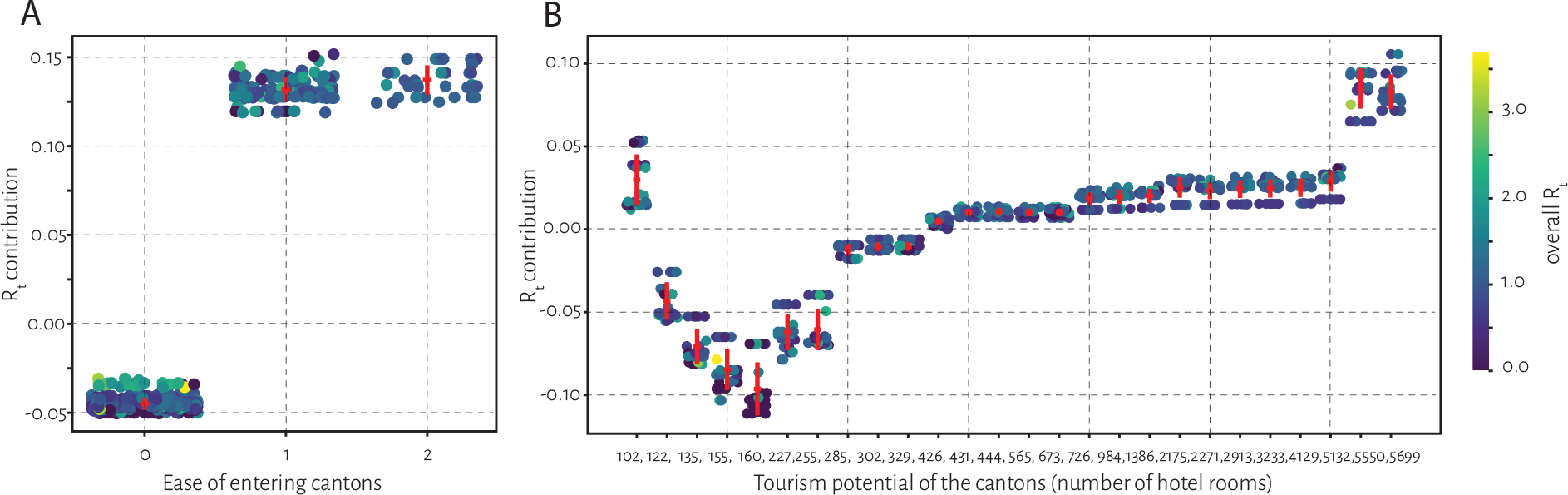
Impact of mobility. The contributions to R_t_ depending on *A*. the ease of entering the cantons by road or air travel where the codes 0 – no travel allowed, 1 – opening of Schengen borders, 2 – opening of international airports with quarantine on travelers were applied and *B*. Tourism potential, represented by the number of hotel rooms in the different cantons are indicated. The non-intuitive higher contributions to R_t_ in cantons with lower tourism potential, can be understood by the borders they share with other Schengen countries. At this moment, because of the lack of access to the actual travel data within Switzerland, it is not easy to infer whether this pattern is due to the commuters or domestic tourism.

#### 2.2.4 Mask requirements

Swiss Federal government made the use of masks in all public transport mandatory (code 1) since July 6. In subsequent weeks, many cantons made the use of a mask to enter the shops (code 2) compulsory. Compared to the no-mask requirement from the pre-lockdown period, a mandatory mask in public transport contributed to a reduction of 0.0139 (CI. 0.0132-0.0144) (Figure 4A). An additional requirement of using the masks in the shops when a requirement in public transport is already mandated (code 2) did not reduce R_t_ further. The use of mask in secondary schools was also made compulsory in some cantons (code 1), and its additional benefit relative to a scenario with schools not requiring masks (code 2) could be seen in Figure 4B. A reduction of R_t_ 0.011 (CI. 0.008-0.0127) was estimated because of the use of the masks in school. The combined effect of the use of mask in public transport and at schools is thus a reduction in R_t_ of 0.025 (CI. 0.018-0.030). It must be noted that the compliance in general may be different from the policy itself, where more people use the mask even if there is no recommendation. This will lead to underestimation of the beneficial effects of the use of the mask.

**Figure 4.**
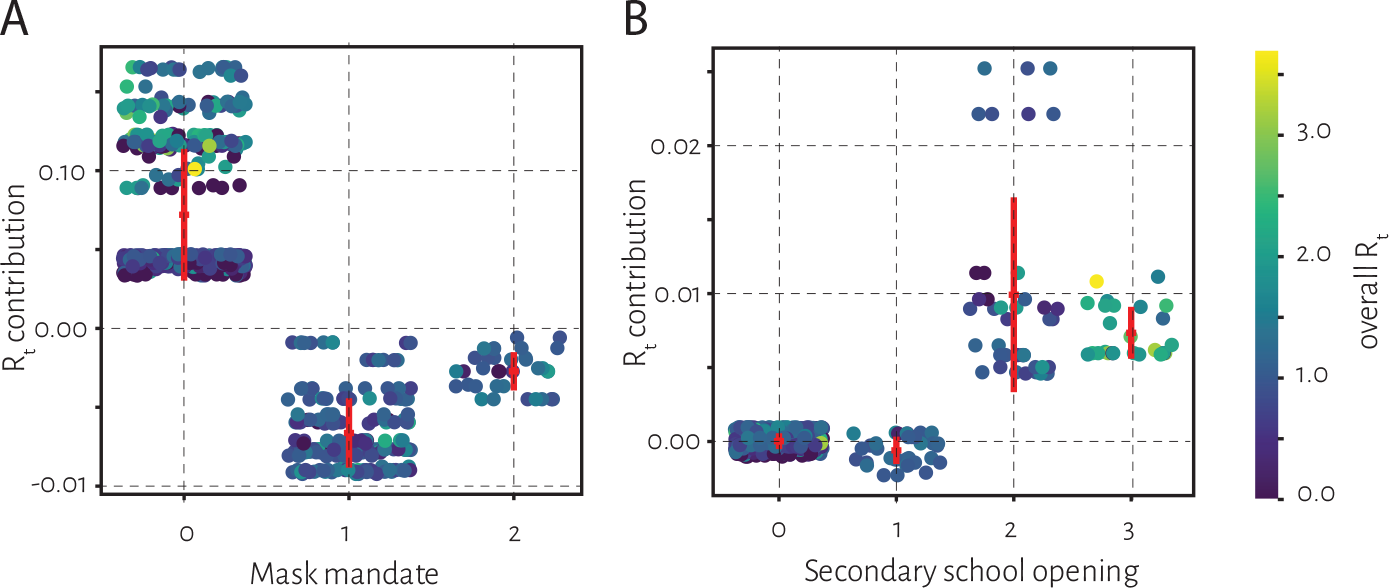
Impact of mask mandates. The mask imposition in *A*. public transport (code 1) and shops (code 2) and *B*. secondary schools (code1), and no-mask restrictions in schools (code 2) are compared. In both the settings the role of imposing a mask is clear.

## 3. Discussion

### 3.1 First estimates for the measure of effectiveness of individual NPIs

Epidemiological scenario calculations forecast the number of infections and critical-care requirements for the various R_t_’s which are in-turn influenced by the NPIs. With an objective to understand how the different NPIs influence the reproduction ratios, the present work addresses the interesting and practically relevant zone between the extremes of a complete lockdown and no restrictions whose role on R_t_ is obvious. The present work attempts to decipher the contribution of each individual NPI implemented at a certain degree of restrictiveness. To the best of our knowledge there has been no precedence on the decomposition of the effectiveness of the different NPIs.

If we consider a mask mandate, there is no evidence for its effectiveness other than referring to the widespread use of masks in the Asian countries where the R_t_’s have been low. The efficacy of masks in reducing aerosol transmission under laboratory conditions [17] are cited as alternative evidences, although their relevance for real scenarios of mask usage in daily life and the impact on the transmission of the disease rather than aerosols is not clear. Our work which interprets what happened in real-life mask usage in public transport and in schools contributed to the reduction R_t_ is a better reflection of how the NPI policies affected COVID-19 transmission. These theoretical inferences for the individual NPI effectiveness are important as it is not easy to design randomized control trials which study each of the individual policies [18].

### 3.2 Overall effectiveness including all implicit transmissions

Switzerland had employed a very efficient contact tracing system [14] which had identified several clusters of infections arising in different business interactions or in social contexts. However, the overall transmissions also include secondary transmissions or via asymptomatic persons, which may not be traced easily. Further, beyond a certain number of daily cases the infrastructure may not support efficient contact tracing. The method we propose here, while may appear redundant when the number of daily infections are few and contact tracing is efficient, will maintain its validity by identifying all potential downstream infections including those by asymptomatics. Although the method will not be helpful in identifying or isolating the infected, it can in principle quantify the impact of NPIs even when the contact-tracing is limited.

### 3.2 Non-causal impact of NPIs

While we underscore the importance of decoupling the contributions using AI in this work, or alternatively by any other model, we also understand that these relations do not necessarily imply causality. The SHAP method identifies the best way of assigning the contributions to the different factors, in each individual prediction, but may not interpreted as causal. Despite this lacuna, the interpretable AI formulation of SHAP designed with the intent of creating “controllable knobs” which can be tuned to achieve the desirable result can be a helpful guide for the “evidence-based” or “data-driven” [19] NPI policies.

### 3.3 Bringing epidemiology and policy-making together using AI

Our work integrates three different perspectives which attempt to understand the mitigation of COVID-19. Policy experts quantify the “stringency” of the Government policies to understand if the response is adequate [20], however, those studies typically do not quantify the consequences of the policies making a discussion on adequacy irrelevant. Epidemiological scenario studies discuss various scenarios of how the critical care requirements are for different possible the reproduction rates, and underscore the importance of maintaining lower reproduction rates without providing a prescription for how it can be achieved. Artificial intelligence has been used to make forecasts, such as predictions of the peaks. However, the predictions hold meaning for a shorter time until either the Government policy or behavioral patterns change. Thus, without addressing the root cause which is the relation between the NPIs and the reproduction ratios, the predictions lose relevance very quickly. In our work we attempt to bring together the policy principles, epidemiological interest in R_t_, within the framework of AI to achieve this evidence for the effectiveness of the different NPI policies.

### 3.4 Learning from data and moving forward

As the “new-normal” with COVID-19 appears to last long, it is important to understand the relative effectiveness of different NPIs for prioritizing them. For example, as per our analysis, and given the behavioral patterns until September, the effectiveness of keeping the borders closed is comparable to the effectiveness of restricting the nightclubs. However, their impacts on the overall society and economy may be completely different and can be weighed in other studies. The model is a first attempt at identifying the contributions of the different NPIs to the R_t_. However, with time more data and considering newer scenarios such as seasonal changes, variable compliances to the NPIs, the model can be trained better. The model should be a living-model, which is regularly updated for reflecting the scenarios better.

## 4. Conclusions

As the COVID-19 pandemic continues to persist for foreseeable future, drastic complete lockdowns may not be a sustainable option. The need for tailored and evidence-based non-pharmaceutical intervention policies which achieve an optimal balance between effectiveness in mitigation and inconvenience in other spheres requires an understanding of the effectiveness of individual policies. The model developed here for deciphering the individual contributions of the various simultaneously imposed non-pharmaceutical intervention policies to the time varying reproduction ratio is a first attempt for such understanding. While the model has to be constantly updated to stay relevant, it paves the possibility for an evidence-based or data-driven policy making where educated, scientifically justifiable and impactful choice of the non-pharmaceutical interventions can be made.

## Data Availability

The analysis is based on publicly available data of number of new infections. The scripts of our analysis are posted in github 
https://github.com/meherpr/SwissRt
and the link is in the manuscript

## Author contributions

CKS, MRB performed the machine learning calculations; MRB, HJ, BS extracted the epidemiological data and performed analyses of rates; CKS, MRB, HJ, BS, MKP analyzed the results; MKP conceived the project, curated the data on government policies and wrote the paper;

## Supplementary Data Availability

The source data, and figures for all states are available at https://github.com/meherpr/SwissRt

